# Establishing syndromic surveillance of gastrointestinal infections in emergency departments using routine emergency department data and validating it against laboratory-based surveillance, Germany, January 2019 – June 2023

**DOI:** 10.1101/2023.11.28.23298985

**Authors:** Jonathan Hans Josef Baum, Achim Dörre, Tamara Sonia Boender, Katharina Heldt, Hendrik Wilking, Susanne Drynda, Bernadett Erdmann, Rupert Grashey, Caroline Grupp, Kirsten Habbinga, Eckard Hamelmann, Amrei Heining, Heike Höger-Schmidt, Clemens Kill, Friedrich Reichert, Joachim Riße, Tobias Schilling, AKTIN Research Group, Madlen Schranz

**Affiliations:** Robert Koch Institute (RKI), Department of Infectious Disease Epidemiology, Berlin, Germany; Postgraduate Training for Applied Epidemiology (PAE), Department of Infectious Disease Epidemiology, Robert Koch Institute, Berlin, Germany; ECDC Fellowship Programme, Field Epidemiology path (EPIET), European Centre for Disease Prevention and Control, (ECDC), Stockholm, Sweden; Charité – Universitätsmedizin Berlin, corporate member of Freie Universität Berlin and Humboldt, Universität zu Berlin, Institute of Public Health, Berlin, Germany; Robert Koch Institute, Risk Communication Unit, Berlin, Germany; Robert Koch Institute, Methods Development, Department of Research Infrastructure and Information Technology, Berlin, Germany; Otto Von Guericke University, Department of Trauma Surgery, Magdeburg, Germany; Klinikum Wolfsburg, Emergency Department, Wolfsburg, Germany; Klinikum Memmingen, Emergency Department, Memmingen, Germany; Ostalb-Klinikum Aalen, Emmergency Department, Aalen, Germany; University Medicine Oldenburg, Pius-Hospital Oldenburg, Clinic for interdisciplinary emergency medicine, Oldenburg, Germany; University Bielefeld, University Hospital OWL, Children’s Center Bethel, Department of Pediatrics, Bielefeld, Germany; Paracelsus Klinikum Henstedt-Ulzburg, Emergency Department, Henstedt-Ulzburg, Germany; Klinikum Chemnitz gGmbH, Emergency Department, Chemnitz, Germany; University Hospital Essen, Center of Emergency Medicine, Essen, Germany; Klinikum Stuttgart, Olgahospital, Interdisciplinary pediatric emergency department PINA, Stuttgart, Germany; Klinikum Stuttgart, Emergency Department, Stuttgart, Germany

## Abstract

2.

**Background:** Gastrointestinal infections in Germany account for 24.5 million outpatient visits annually. Surveillance of gastrointestinal infections in emergency departments strengthens timely outbreak detection and disease trend monitoring.

**Aim:** We developed a syndrome definition for automated syndromic surveillance of gastrointestinal infections in emergency departments, and validated it against statutory laboratory-based surveillance.

**Methods:** To develop a syndrome definition, we selected presenting complaints (Canadian Emergency Department Information System) and diagnoses (ICD-10). We validated the definition through time series and cross-correlation analysis, comparing trends between syndromic and laboratory-based surveillance. We analysed German emergency department registry (AKTIN) data and included emergency departments that continuously transferred (01/2019-06/2023) data. As reference we combined statutory norovirus-gastroenteritis, rotavirus-gastroenteritis, campylobacteriosis and salmonellosis notifications.

**Results:** Our syndrome definition combined presenting complaints (diarrhoea, vomiting and nausea) and diagnoses (Intestinal infectious diseases). Accordingly, in 7 emergency departments with *n* = 864,353 visits, 2.1% (*n* = 18,158) were gastrointestinal infection cases. Of those, 57% (*n* = 10,424) were female, with 23% 0–19 years (*n* = 4,108) and 23% 20–29 years (*n* = 4,116) old. We visually observed similar gastrointestinal infection trends in both surveillance systems. The cross-correlation was 0.73 (95%-confidence interval 0.61–0.85; *p*<0.001) at lag −1, indicating a 1-week relative reporting delay of laboratory-based surveillance.

**Conclusion:** The coherent trends and significant cross-correlation validated our syndrome definition, which adequately captures gastrointestinal infection cases in emergency departments. Our novel automated surveillance complements laboratory-based surveillance, while offering advantages regarding timeliness and reduced workload. Therefore, it will be implemented in national routine surveillance.

## 3. Introduction

Gastrointestinal infections are of viral, bacterial, fungal or parasitic aetiology and cause gastroenteritis which is an inflammation of stomach and intestine. The main symptoms are nausea, vomiting and diarrhoea (1). These infections are among the top eight causes of death globally (2), and are a considerable burden on health care services. In 2008 and 2009, acute gastrointestinal illness was estimated to cause about 24.5 million outpatient visits and 19.9 million hospital days per year in the adult population in Germany (3). Norovirus-gastroenteritis, campylobacteriosis, and rotavirus-gastroenteritis ranked among the top four in incidence of notifiable diseases in Germany in 2019 prior to the COVID-19 pandemic (4). Surveillance of gastrointestinal infections is important to monitor disease trends and identify gastrointestinal outbreaks. It helps to assess the public health impact and better understand disease mechanisms and causes. Thereby surveillance improves prevention and outbreak detection to enable timely public health action.

Data on the occurrence of gastrointestinal infectious diseases in Germany is collected through the statutory surveillance system, where the majority of gastrointestinal infectious disease notifications are laboratory confirmed. Occasionally, some notifications are included based on clinical presentation and an epidemiological link only. Importantly, this epidemiological link is again to another laboratory-confirmed case (5). Although notifiable, gastrointestinal outbreaks are often not reported. Laboratory diagnostics and reporting delays cause a time lag. The existing system could be complemented by establishing syndromic surveillance, which is not based on laboratory diagnoses, but on clinical signs and symptoms that constitute a “syndrome” (6).

Emergency departments are suited for syndromic surveillance. Firstly, they routinely collect symptom-based information such as presenting complaints or preliminary diagnosis. Secondly, the specific setting of emergency departments is highly relevant for surveillance, since emergency departments often serve as the first point of contact for patients with the healthcare system. Surveillance in emergency departments could support the description of trends and timely detection of outbreaks, particularly regarding more severe gastrointestinal infections. Additionally, the reduced specificity of syndromic surveillance may increase sensitivity for the detection of unspecific, yet relevant public health events.

In Germany, syndromic emergency department surveillance was established at the Robert Koch Institute in 2018. The system uses routinely collected case-based data provided daily by the AKTIN emergency department registry (7, 8). Based thereupon, the Robert Koch Institute publishes an automated weekly emergency department report (9), including syndromes such as (severe) acute respiratory infection or influenza-like illness (10). Gastrointestinal infections are not yet included in routine reporting. Preliminary syndrome definitions for gastrointestinal infections were composed in an earlier project, but required validation and further adaptation for the implementation in surveillance reporting (11).

In order to describe disease trends and facilitate timely outbreak detection, we aimed to develop and implement syndromic surveillance of gastrointestinal infections in emergency departments in Germany. Therefore, our objectives were to further develop a syndrome definition for gastrointestinal infections, to describe the distribution of identified cases over time and to validate the syndrome definition by comparison with laboratory-based reference data.

## 4. Methods

### Development of a syndrome definition for gastrointestinal infections

We combined both ICD-10 coded diagnosis and presenting complaints to develop a syndrome definition (case definition) for gastrointestinal infections. It was based on syndrome definitions developed in a previous study (11). The selection of relevant ICD-10 diagnosis and presenting complaints was inspired by similar syndrome definitions published in the literature (12, 13) and developed in consultation with the unit for gastrointestinal infections at the Robert Koch Institute. It was further discussed and refined in collaboration with the AKTIN Research Group and clinicians from participating emergency departments.

### Study data bases

We performed a retrospective registry study using data provided by the AKTIN emergency department registry (7, 8). AKTIN provides the emergency department surveillance system at the Robert Koch Institute with case-based routine emergency department data on a daily basis (AKTIN data request ID2019_003). The data is then transformed according to the NoKeDa (NotaufnahmeKernDatensatz) data standard (14) and stored in a local database at the Robert Koch Institute. Participation in the AKTIN emergency department registry is voluntary for emergency departments, which means that the sample is non-comprehensive and not necessarily geographically representative. Each record in the dataset corresponds to one patient visit to the emergency department, described by a unique identifier. Since the data is anonymized, multiple visits for the same patient cannot be linked.

We included the following variables in our analysis: emergency department, date of visit, sex, age (in age groups 0–19, 20–39, 40–59, 60–79, 80+ years), presenting complaints (according to the Canadian Emergency Department Information System – Presenting Complaint List V.3.0 (15)), diagnosis coded according to the German modification of the 10^th^ International Statistical Classification of Diseases and Related Health Problems (ICD-10) and ICD-10 diagnosis certainty (16). We removed the relatively rare sex category “other” to ensure data protection and patient privacy in all analysis. We exported data from the database on 1 August 2023.

For the laboratory-based reference, we combined the number of notifications of norovirus-gastroenteritis, rotavirus-gastroenteritis, campylobacteriosis, and salmonellosis from the German statutory electronic surveillance system (SurvNet) by week of notification. We included notifications fulfilling case definitions according to §11 (2) Infection Protection Act (5). We did not include other notifiable gastrointestinal diseases (e.g. giardiasis, shigellosis) due to relatively small case numbers. SurvNet data was exported on 20 August 2023.

### Study period, study population and selection of emergency departments

We defined the study period as ranging from calendar week 2/2019 (7 January 2019) to week 25/2023 (25 June 2023), in which data from a total of 43 emergency departments across 10 German federal states was available. Of those, we selected departments that fulfilled pre-defined quality criteria. Firstly, we assessed the continuous transmission of data, which was defined as a maximum of one week without any transmitted patient visits during the study period. Secondly, we considered the quality of two main variables, presenting complaint and ICD-10 diagnosis. We allowed a maximum of one week with 100% missing values and no trend for each of the two variables during the study period. We accepted emergency departments who consistently reported only one of the two variables (i.e. one variable continuously missing). In this case the syndrome was consistently coded using only one variable. Data from one emergency department was excluded due to additional technical problems with data quality. This strict quality control reduces fluctuations in our indicator due to variations in data quality, which could distort the subsequent absolute trend and cross-correlation analysis. The study population includes all patients that visited the selected emergency departments in the study period.

### Statistical analysis, internal and external validation of syndrome definition

Using frequency measures, we described emergency department visits as well as cases of gastrointestinal infections by sex and age groups. We described weekly cases of gastrointestinal infections over time and analysed trends and seasonal patterns through time series analysis.

To internally validate the use of presenting complaints for the selection of gastrointestinal infection cases, we performed an additional analysis. We selected cases solely based on presenting complaints and subsequently analysed ICD-10 diagnosis frequencies and consistency with gastrointestinal infections.

For external validation of the syndrome definition, we assessed the correlation between cases of gastrointestinal infections in emergency departments (“emergency department indicator”) and data for gastrointestinal infectious diseases notifications from laboratory-based surveillance (“reference indicator”). First, we visually compared the trends of both the emergency department indicator and the reference indicator. We then quantified the correlation by estimating cross-correlation coefficients between the two indicators, both overall and stratified by sex and age groups. We estimated the standard error with the moving block bootstrap method (17). We applied Loess smoothing for estimating the trend and the corresponding remainder components of each time series as a basis for the block bootstrap, and we defined blocks of length 17 each. Based on an approximation with Student’s *t-*distribution, we calculated confidence intervals and statistical significance (*p*-values) for cross-correlation coefficients at specific time lags.

We used the statistical software R (Version 4.2.2) (18) and RStudio (Version 2023.03.0 Build 386) (19), including the packages *tidyverse*, *lubridate*, *ISOweek*, *Readxl*, *naniar*, *flextable*, *gtsummary*, *stringr*, *tsibble*, *slider*, *ggh4x*, *ggpubr*, *tseries*.

## 5. Results

### Syndrome definition for gastrointestinal infections

The syndrome definition we developed combined presenting complaints “254 – Diarrhea” and “257 – Nausea and/or vomiting” with the ICD-10 diagnosis group “A00 – A09 Intestinal infectious diseases” (***Figure 1***). Visits were defined as cases, if they were coded with any of the respective presenting complaints or ICD-10 diagnosis. For ICD-10 diagnosis, we accepted the diagnoses with certainty levels of “G” (confirmed), “V” (suspected), “Z” (previous) or missing certainty level, but not “A” (excluded).

**Figure 1:**
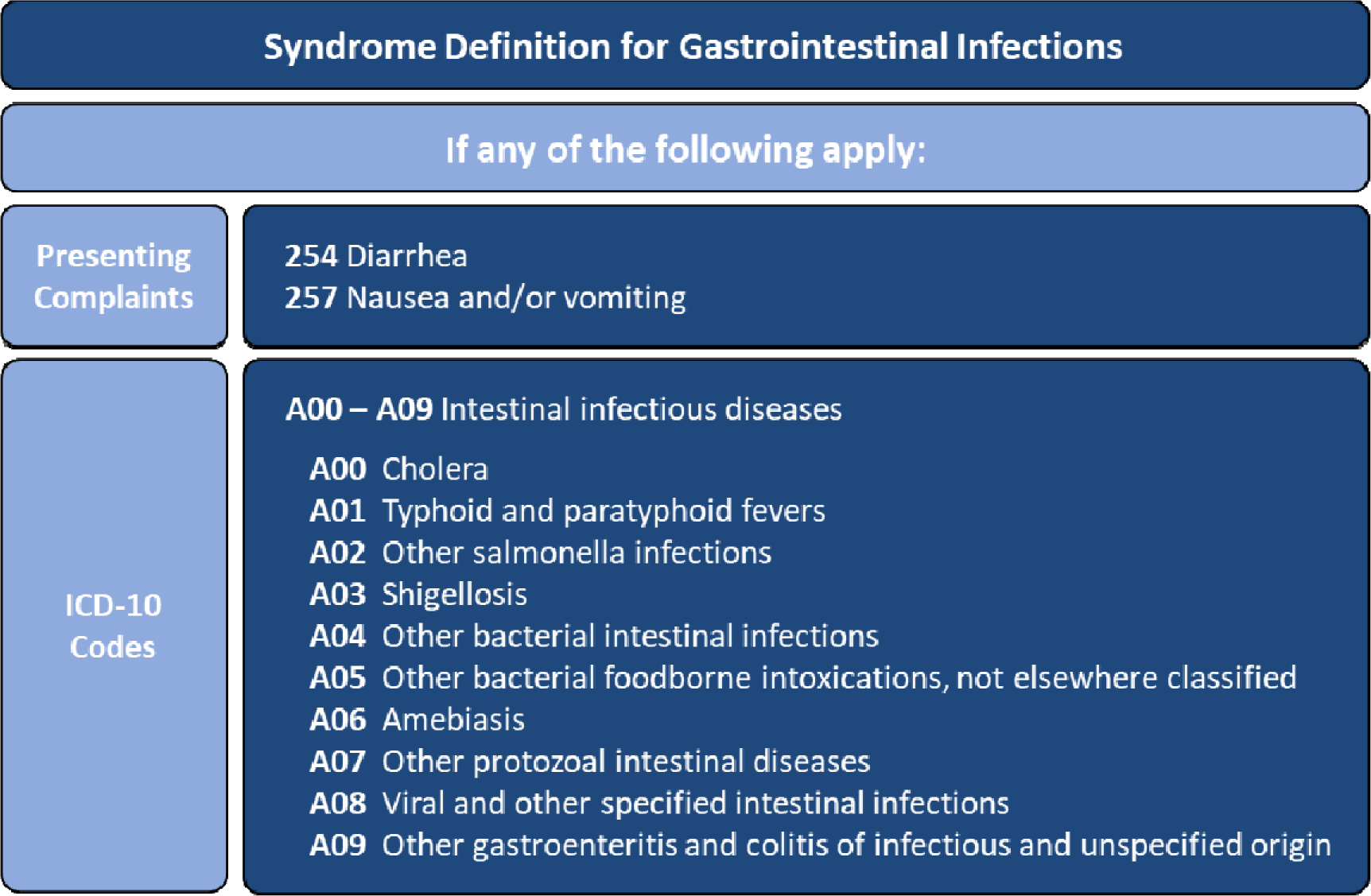
Syndrome definition used for surveillance of gastrointestinal infections in emergency departments by presenting complaint according to the Canadian Emergency Department Information System and ICD-10 codes. Germany, 2019–2023.

Based expert discussions on clinical coding practices, the syndrome definitions for “GI unspecific without bleeding” and “bloody diarrhoea” developed in a previous project (11), were considered not suitable for routine surveillance. First, they included presenting complaints based on the Manchester Triage System, which is currently not included in the data provided by the AKTIN registry. Secondly, the majority of cases of “bloody diarrhoea” received ICD-10 codes “K92.1 Melaena” or “K92.2 Gastrointestinal haemorrhage, unspecified”, which were considered too unspecific for gastrointestinal infections. For the same reason, the presenting complaints “251 – Abdominal pain” or “260 – Blood in stool/melena” were not included in the syndrome definition, nor ICD-10 diagnoses such as “R11 – Nausea and vomiting” or “K92 – Other diseases of digestive system”.

### Selected emergency departments

Within the study period, data from 7 out of 43 emergency departments fulfilled the inclusion criteria. The departments were located in 6 out of 16 German federal states, namely Baden-Wurttemberg, Bavaria, Lower Saxony, North Rhine-Westphalia, Saxony and Schleswig-Holstein. The emergency departments differed greatly regarding patient capacity, with the largest reporting an average of 674 weekly visits (range: 420–829), three times more than the smallest with 216 weekly visits on average (range: 115–284).

### Characteristics of emergency department visits and cases of gastrointestinal infection cases

The 7 included emergency departments registered a total of *n* = 864,353 patient visits during the study period or 3,710 visits per week on average (range: 2,413–4,906 visits per week). Patients aged 20–39 and 60–79 years made up the largest age groups with 24% each of the visits. 51% of visits were male. According to the syndrome definition, we classified 2.1% (*n* = 18,158) of visits as cases of gastrointestinal infections, ranging between 0.8% and 4.3% among emergency departments. Most cases of gastrointestinal infections were either 0–19 years or 20–29 years old with 23% each. 57% of cases were female (***Table 1***).

**Table 1:**
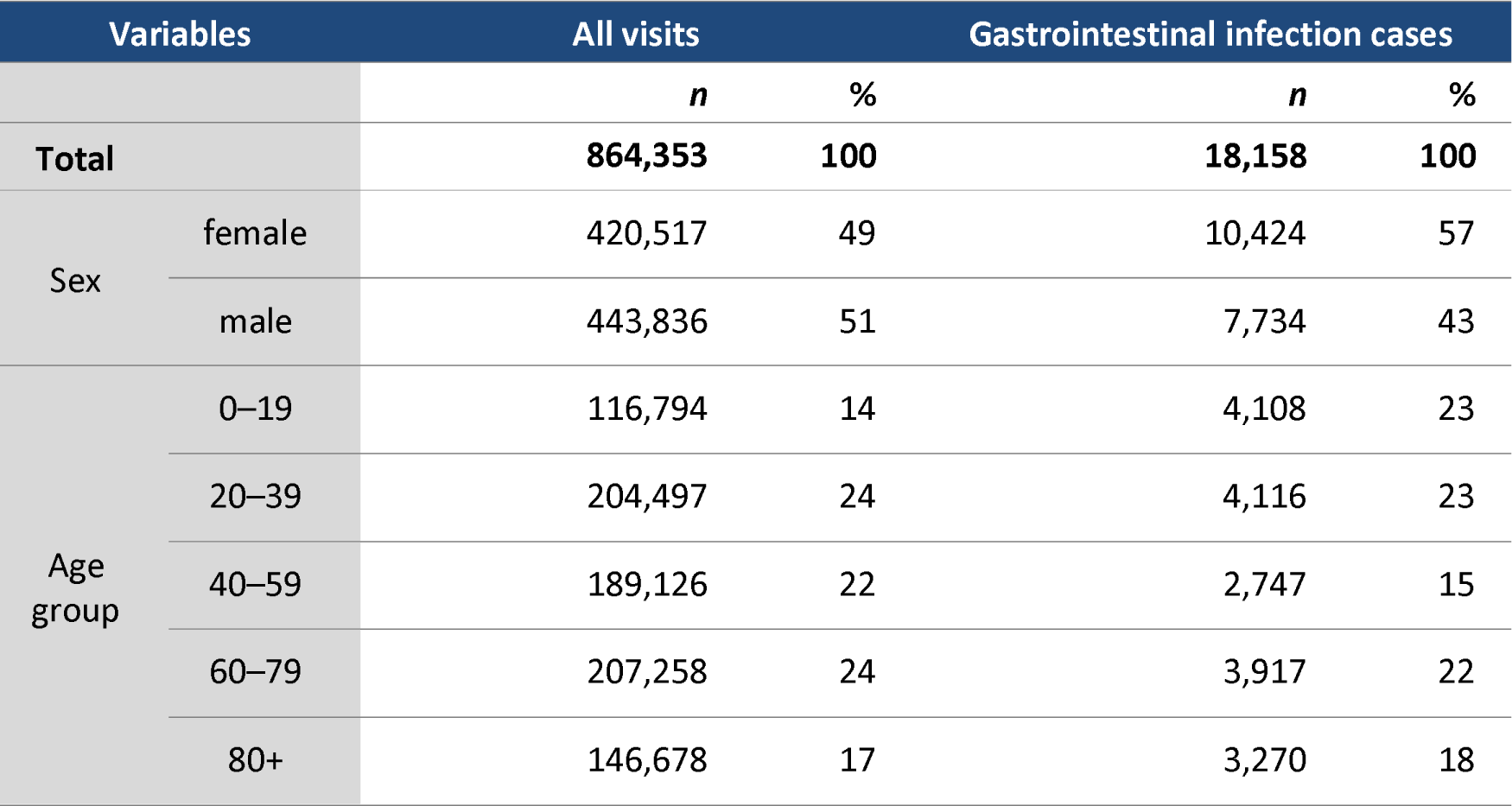
Characteristics of emergency department visits and cases of gastrointestinal infection cases. Germany, 2019–2023.

### Internal validation of the syndrome definition

To assess internal validity of presenting complaints, we selected cases solely through the two presenting complaints “254 – Diarrhea” and “257 – Nausea and/or vomiting”. We subsequently analysed the frequencies of ICD-10 codes, of which the following dominated, all consistent with gastrointestinal infections: Diagnoses “A09.9 – Other and unspecified gastroenteritis and colitis of unspecified origin”, “A09.0 – Other and unspecified gastroenteritis and colitis of infectious origin”, and “R11 – Nausea and vomiting”. Conversely, we observed few ICD-10 codes inconsistent with gastrointestinal infections (***Supplementary table 1*** and ***Supplementary table 2***).

**Supplementary table 1:** Frequency of ICD-10 codes in visits (n = 10,352) selected based on presenting complaint “254 – Diarrhea”. ICD-10 codes with frequencies ≤1% were not shown. Frequencies were calculated relative to all visits with available ICD-10 codes. Germany, 2019 – 2023.

| ICD-10 codes |  | n | Frequency % |
| --- | --- | --- | --- |
| **A09.9** | Other and unspecified gastroenteritis and colitis of unspecified origin | 3,093 | 29.9 |
| **E86** | Volume depletion | 504 | 4.9 |
| **A09.0** | Other and unspecified gastroenteritis and colitis of infectious origin | 503 | 4.9 |
| **R11** | Nausea and vomiting | 334 | 3.2 |
| **Z29.0** | Isolation as prophylactic measure | 310 | 3.0 |
| **Z11** | Special screening examination for infectious and parasitic diseases | 206 | 2.0 |
| **R10.4** | Other and unspecified abdominal pain | 187 | 1.8 |
| **R53** | Malaise and fatigue | 185 | 1.8 |
| **E87.1** | Hypo-osmolality and hyponatraemia | 127 | 1.2 |
| **E87.6** | Hypokalaemia | 125 | 1.2 |
| **N17.9** | Acute renal failure, unspecified | 111 | 1.1 |
| **N39.0** | Urinary tract infection, unspecified | 107 | 1.0 |
| **K52.9** | Noninfective gastroenteritis and colitis, unspecified | 106 | 1.0 |

**Supplementary table 2:** Frequency of ICD-10 codes for visits (n = 10,696) selected based on presenting complaint “257 – Nausea and/or vomiting”. ICD-10 codes with frequencies ≤1% were not shown. Frequencies were calculated relative to all visits with available ICD-10 codes. Germany, 2019 – 2023.

| ICD-10 codes |  | *n* | Frequency % |
| --- | --- | --- | --- |
| **R11** | Nausea and vomiting | 2,194 | 20.5 |
| **A09.9** | Other and unspecified gastroenteritis and colitis of unspecified origin | 867 | 8.1 |
| **R10.4** | Other and unspecified abdominal pain | 308 | 2.9 |
| **R53** | Malaise and fatigue | 229 | 2.1 |
| **E86** | Volume depletion | 224 | 2.1 |
| **R10.1** | Pain localized to upper abdomen | 192 | 1.8 |
| **A09.0** | Other and unspecified gastroenteritis and colitis of infectious origin | 179 | 1.7 |
| **R42** | Dizziness and giddiness | 152 | 1.4 |
| **K92.0** | Haematemesis | 144 | 1.3 |
| **Z11** | Special screening examination for infectious and parasitic diseases | 141 | 1.3 |
| **N39.0** | Urinary tract infection, unspecified | 128 | 1.2 |
| **E87.1** | Hypo-osmolality and hyponatraemia | 122 | 1.1 |

### External validation of the syndrome definition

We obtained the laboratory-based reference indicator by combining statutory notifications of norovirus-gastroenteritis, rotavirus-gastroenteritis, salmonellosis and campylobacteriosis (***Supplementary figure 1***). For the visual comparison of trends of gastrointestinal infections over time, we plotted the emergency department indicator and the reference indicator by calendar week (***Figure 2***). Both indicators exhibited similar trends. Especially, a marked decrease in the first half of 2020, followed by a slight increase over the summer 2020 and another decrease in Winter 2020/21 was apparent for emergency department and reference indicator alike. For both indicators, the number of cases rose again towards summer 2021, followed by peaks in summer 2022 and spring 2023.

**Supplementary figure 1:**
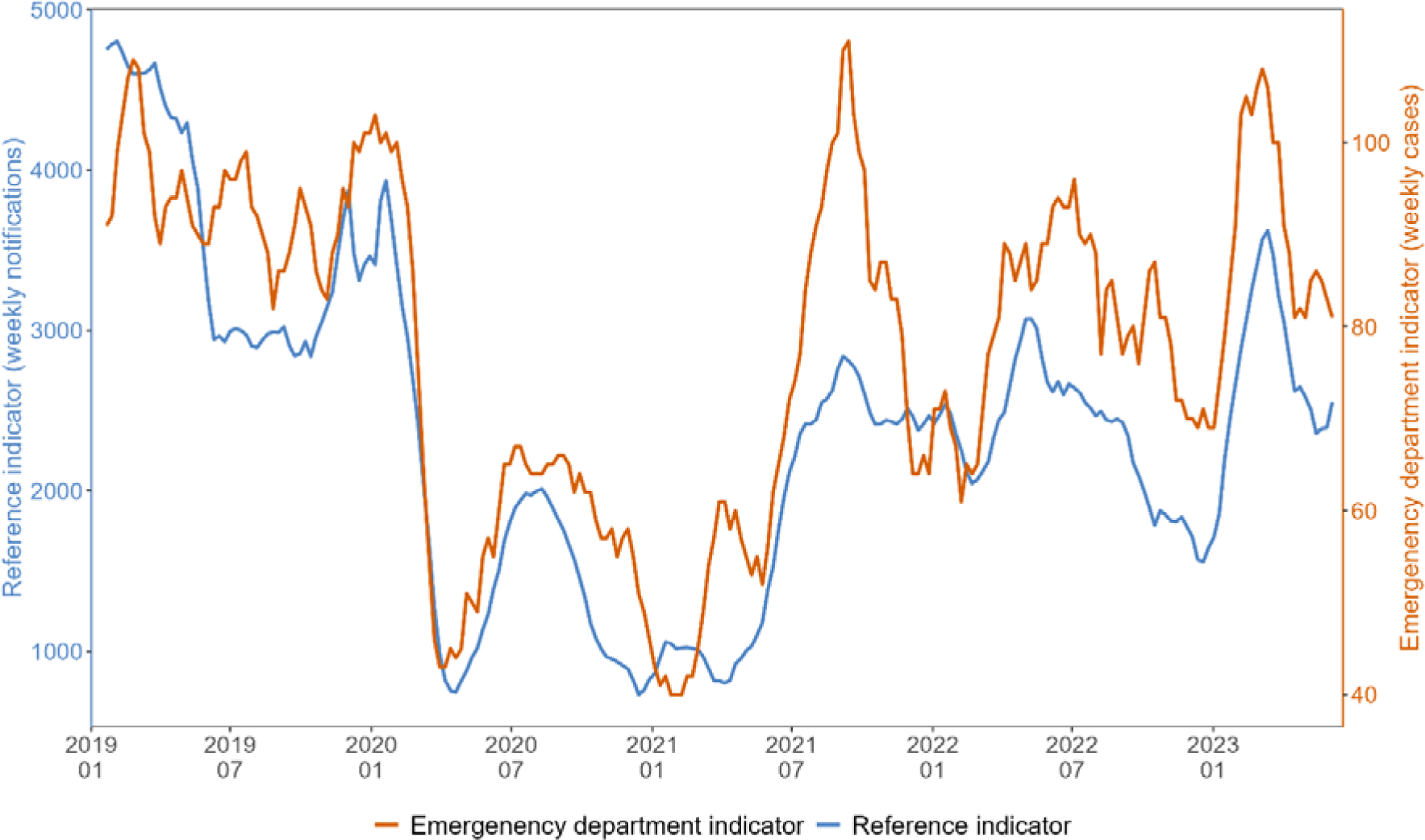
Notifications for norovirus-gastroenteritis, rotavirus-gastroenteritis, salmonellosis and campylobacteriosis from laboratory-based surveillance and derived combined reference indicator. Germany, between week 01/2019 and up to week 23/2023.

**Figure 2:**
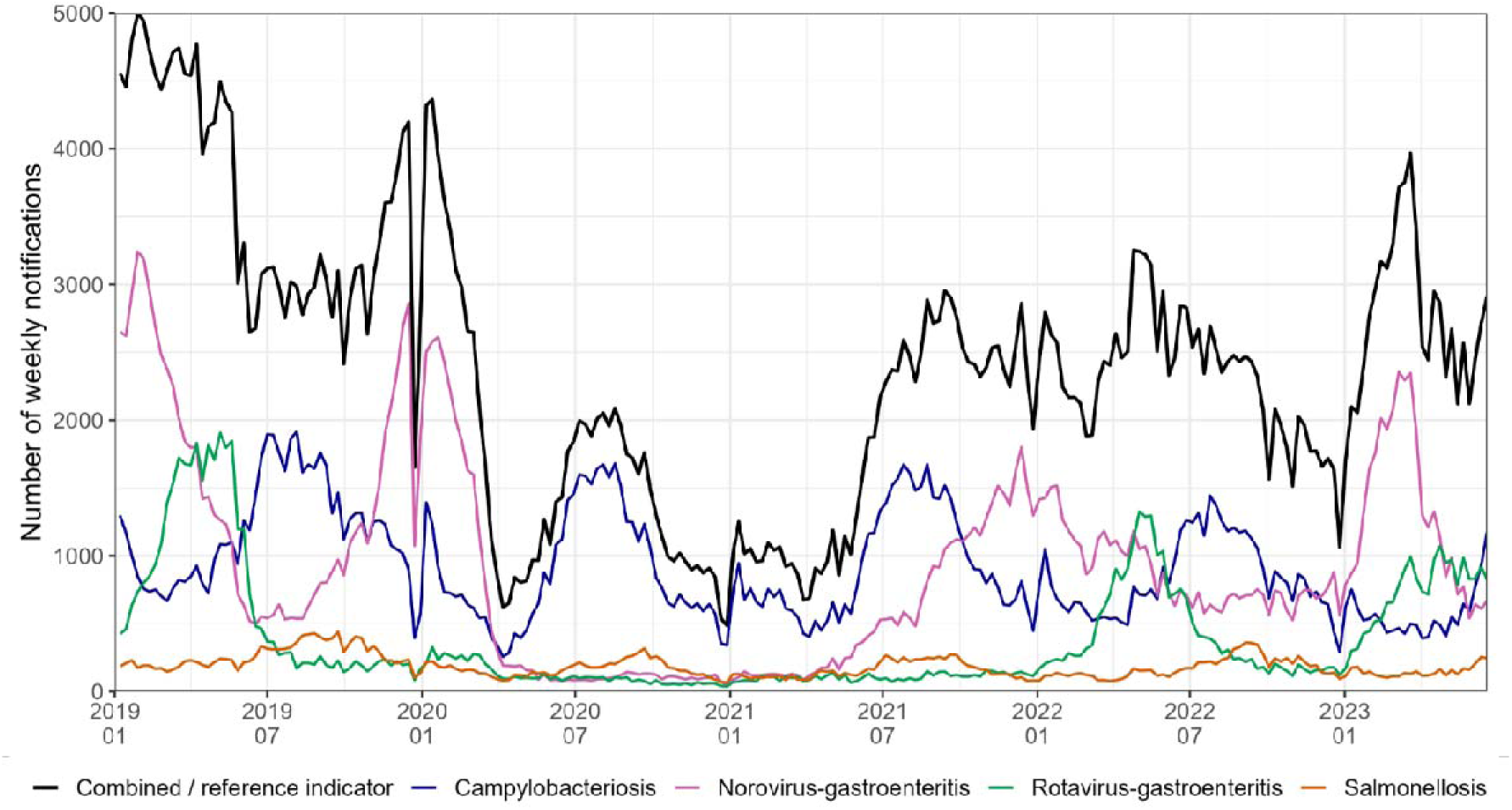
Visual comparison of emergency department indicator (orange) and combined reference indicator (blue) trends as 5-week-moving-averages. Please note different scales on left and right y-axes. Germany, between week 01/2019 and up to week 23/2023.

Upon stratification by age groups, younger age groups showed high coherence between the trends of both indicators. For older age groups, especially above 80 years of age, coherence between the two indicators was relatively lower than overall (***Figure 3***).Cross-correlation analysis between the two indicators showed the highest correlation at a time lag of −1 (in weeks), indicating that gastrointestinal infection trends in emergency departments were one week ahead of reference notification data. The overall cross-correlation coefficient at lag −1 was 0.73 (95%-confidence interval 0.61–0.85; *p*<0.001). Cross-correlation analysis by sex and by age at lag −1 revealed high correlation in all strata, with a lower value for patients older than 80 years of age (***Table 2***).

**Figure 3:**
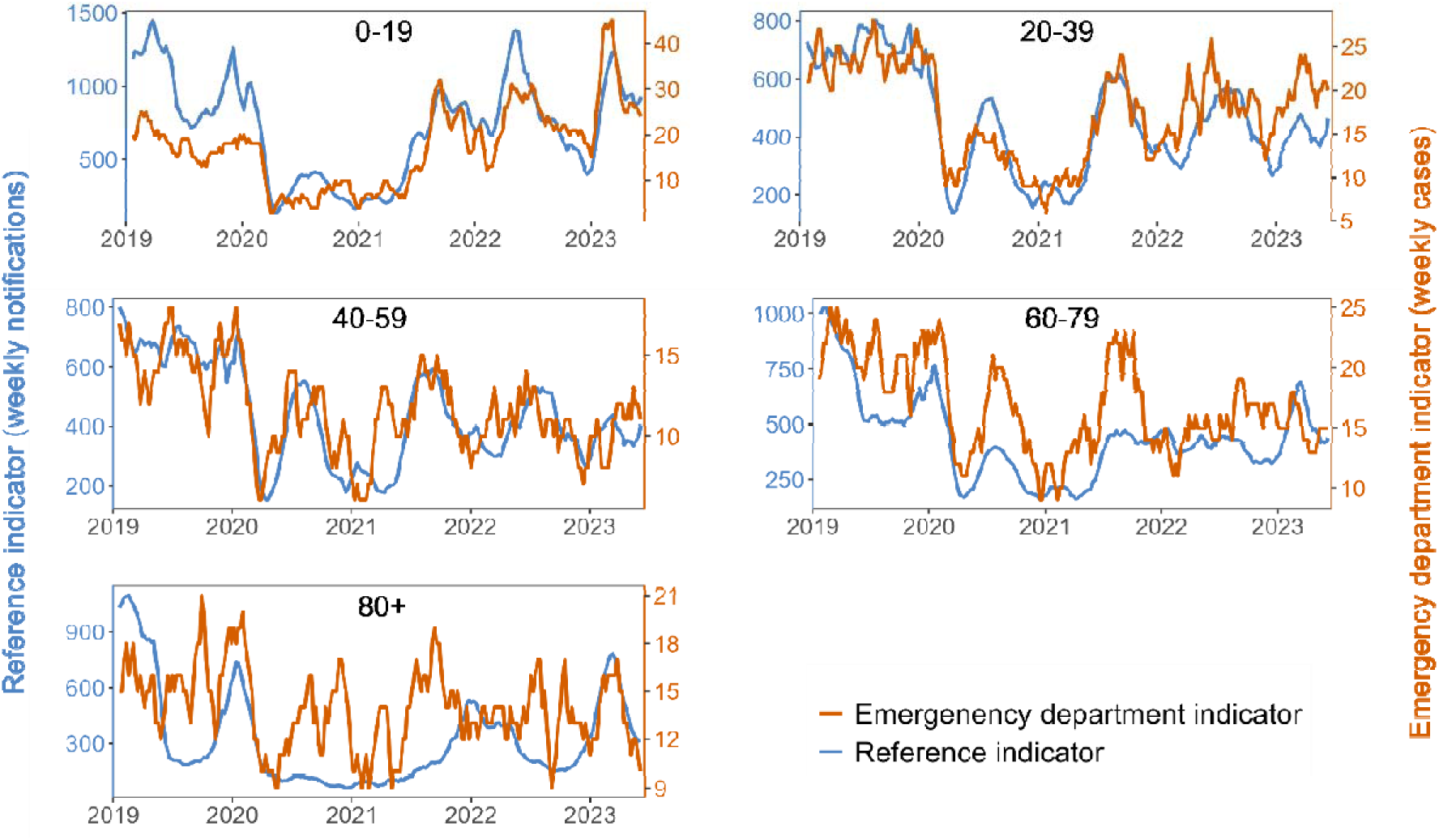
Visual comparison of emergency department indicator (orange) and reference indicator (blue) trends as 5-week-moving-averages stratified by age groups. Please not different scales on left and right y-axis. Germany, between week 01/2019 and up to week 23/2023.

**Table 2:** Results of emergency department indicator and reference indicator cross-correlation analyses at lag −1 overall, and stratified by sex and age groups. *P-values refer to the statistical significance of the cross-correlation coefficient being different from 0. Germany, 2019–2023.

|  |  | Cross-correlation coefficient | 95% confidence interval | p-value* |
| --- | --- | --- | --- | --- |
| Overall | | 0.73 | 0.61–0.85 | $p<0.001$ |
| Sex | male | 0.67 | 0.56–0.79 | $p<0.001$ |
| | female | 0.65 | 0.50–0.80 | $p<0.001$ |
| Age groups (years) | 0–19 | 0.65 | 0.53–0.77 | $p<0.001$ |
| | 20–39 | 0.61 | 0.47–0.74 | $p<0.001$ |
| | 40–59 | 0.48 | 0.35–0.61 | $p<0.001$ |
| | 60–79 | 0.48 | 0.33–0.63 | $p<0.001$ |
| | 80+ | 0.22 | 0.03–0.40 | $p<0.05$ |

## 6. Discussion

We developed a syndrome definition for gastrointestinal infections in emergency departments, which combines presenting complaints (“254 – Diarrhea”, “257 – Nausea and/or vomiting”) and ICD-10 diagnoses (A00-A09: Intestinal infectious diseases). The observed coherent trends and high cross-correlation between our emergency department indicator and laboratory-based reference indicator validated it.

We are confident that our syndromic surveillance will complement laboratory-based surveillance and support the detection of medium and large gastrointestinal outbreaks. For example, in a 2012 outbreak of gastroenteritis associated with the consumption of frozen strawberries, approximately 11,000 cases were detected via ad hoc syndromic surveillance using spreadsheets and emails (23). Our system would be able to detect such outbreaks faster, provide more detailed estimates on severe cases, and simplify outbreak surveillance.

Regarding timeliness, we found the highest overall correlation at lag −1, indicating a 1-week reporting delay for laboratory-based surveillance relative to syndromic surveillance. Compared to existing systems with time-consuming laboratory testing and reporting delays, our automated syndromic surveillance of routine data offers near real-time monitoring. This improves the timely detection of disease trends and outbreaks, thereby facilitating a faster and more effective public health response.

The syndromic surveillance of gastrointestinal infections in emergency departments contributes to an all-hazards surveillance approach. Some unexpected natural or man-made health events may be missed by laboratory-based and pathogen-specific surveillance due to unavailable or delayed laboratory test results. Syndromic surveillance, on the other hand, may identify these events earlier, based on an increase of patients with respective symptoms in emergency departments. For example, the system may support the detection of (accidental or intentional) chemically-induced gastroenteritis or events of bioterroristic origin.

By combining routine data from several sentinel emergency departments in multiple federal German states, the system supports the identification of superregional gastrointestinal disease trends and outbreaks. Moreover, our syndromic surveillance enhances real-time situational awareness during special events such as mass gatherings or sports events. In 2024, our syndromic surveillance for gastrointestinal infections will help to monitor a mass sporting event in Germany, the UEFA European Football Championship.

Compared to laboratory-based surveillance, the system offers a reduced workload due to its automated nature. Moreover, since it is based on routinely collected data and the existing AKTIN-infrastructure, it is comparably inexpensive. The use of an existing system, whose infrastructure matured and operated since 2018, ensures the stability and resilience of the gastrointestinal infection surveillance component.

Our syndrome definition combines two syndromic components. The selected ICD-10 diagnoses are more specific for gastrointestinal infections, but may be coded with a delay. Presenting complaints are coded directly upon admission and prior to detailed anamnesis. Therefore, they provide near real-time insights, but may lack diagnostic precision. In our analysis of the internal validity of presenting complaints, we observed few ICD-10 codes inconsistent with gastrointestinal infections. Integrating both data sources thus creates a suitable compromise between sensitivity and specificity to facilitate accurate and timely surveillance.

Visual trend comparison showed high coherence of trends of emergency department and reference indicators, further supported by the high cross-correlation. Observed separately, the selected reference diseases showed characteristic seasonality and peaks, albeit partially suspended during the COVID-19 pandemic (***Supplementary figure 1***). Such seasonality was not observed in either indicator, due to several diseases with shifted seasonality overlapping. This highlights that our emergency department indicator is not dominated by one disease, but rather reflects an ensemble of gastrointestinal diseases. We observed a decrease during the COVID-19 pandemic in both indicators. A similar decrease was also observed in other syndromic surveillance systems for gastrointestinal complaints at the same time, for example in the United Kingdom (20).

Cross-correlation analysis corroborated the visual results with a high overall coefficient of 0.73 at lag −1. Upon stratification by sex and age groups, we found high visual coherence in most strata, supported by high cross-correlation coefficients. Notably, cross-correlation was lower in patients older than 80 years old. A possible explanation could be differences in clinical presentations of gastrointestinal infections in older patients or age-specific testing and coding practices. The number of weekly patients in this stratum was also relatively low, rendering the indicator more susceptible to fluctuations. Moreover, concurrent COVID-19 waves may have resulted in higher symptomatic gastrointestinal presentations in emergency departments, which are absent in gastrointestinal notifications. Diarrhoea has been linked to COVID-19, particularly in older patients (21). Changed health-seeking behaviour may have resulted in older patients having a higher probability of admission to emergency departments due to higher risk for severe COVID-19. Potentially, this limits our ability to detect some smaller events in age-specific settings. However, syndromic surveillance does not aim to identify every small event, but rather to describe overall trends and detect events with high public health relevance.

The highest proportions of identified cases of syndromic gastrointestinal infections were among females and patients aged 39 and below. Similarly, women and younger age groups were also overrepresented in studies on gastroenteritis, e.g. from general practice in Norway (22) and US emergency departments (23). We identified 2.1% of all visits as cases of gastrointestinal infection, according to our syndrome definition. To compare this to the literature, a 2017 study of 5 German emergency departments found 12% of patients presented symptoms related to the digestive system (24). In the weekly reports of the Italian national emergency department surveillance for 2014, the percentage of patients with non-haemorrhagic gastroenteritis fluctuated between 1.3–2.3% (25). Comparisons with other studies and syndromic surveillance systems are limited by differences in case definitions, clinical settings and demographics.

In Germany, no syndromic surveillance of gastrointestinal infections is currently operational. An ad-hoc surveillance for bloody diarrhoea during an outbreak of enterohemorrhagic *E. coli* was established in 2011, but later discontinued. Syndromic surveillance systems are increasingly implemented in several European countries. Since 2004 France has a surveillance system with national coverage. It includes gastroenteritis and covers ≥86% of emergency department visits. The United Kingdom developed subnational systems, which regularly publish weekly bulletins including gastroenteritis in emergency departments. In Italy, national and regional systems in Genoa and the Lazio region include syndromes for non- and haemorrhagic gastroenteritis. Similar systems were implemented in selected hospitals in Austria and Spain. Greece established a sentinel surveillance system for the 2004 Olympics (12, 26–28). Syndromic surveillance systems proved useful in the early identification of gastrointestinal disease trends and outbreaks, e.g. of rotavirus-gastroenteritis (29) and enterohemorrhagic *E. coli* (28) and for assessing potential health impacts of mass gatherings (30).

Our findings should be interpreted considering some limitations. As a sentinel surveillance system, the AKTIN registry is neither geographically nor demographically representative for the German population. Data collected through routine hospital documentation is primarily intended for clinical use rather than disease surveillance. The quality of documentation may vary between emergency departments, resulting in possible misclassification bias. Particularly, the coding of diagnoses according to ICD-10 may differ due to (hospital-specific) administrative and budgetary coding regulations. Since syndromic surveillance is not based on laboratory-confirmed diagnoses but on less specific clinical symptoms, cases may be misclassified, possibly resulting in over- or underestimation of case numbers. However, since syndromic surveillance aims at identifying overall trends rather than identifying single cases, it may tolerate some deviation of the estimate. All visit data is anonymised, so there could be multiple visits by the same patient, possibly resulting in excess cases. Nevertheless, as this imprecision effect remains over time, the expected bias is likewise relatively constant over time. Our analysis was complicated by the COVID-19 pandemic coinciding with our study period. Through multiple factors, foremost changes in health-seeking behaviour, the pandemic may have shifted the composition and numbers of patients attending emergency departments in general and cases of gastrointestinal infections specifically (31, 32).

In the future, syndromic surveillance for gastrointestinal infections may be expanded. For example, the system could be adapted for the early identification of relevant severe gastrointestinal conditions, such as “bloody diarrhoea” or “haemolytic uraemic syndrome”. More variables could be added to further refine the syndrome definition, such as body temperature or patient isolation. Signal algorithms could analyse data to trigger automated alerts. As of January 2022, the ICD-11 version officially replaces its ICD-10 predecessor. ICD-11 codes are not yet used by the AKTIN emergency departments. Following a transitional period and their implementation, the syndrome definition may need to be adapted accordingly. The ICD-11 revisions may prove useful, as the new group “1A0 – 1A40 Gastroenteritis or colitis of infectious origin” combines many relevant diagnoses (33). Our analysis was based on retrospective data, but eventually the performance of the system should be evaluated prospectively.

We conclude that the syndrome definition for gastrointestinal infections in emergency departments is valid and ready for implementation in routine surveillance. It will be implemented in the Robert Koch Institute’s automated emergency department surveillance reporting with the next update (planned for end of 2023). Our novel automated surveillance offers advantages regarding timeliness and reduced workload compared to laboratory-based surveillance. It provides an added public health value by completing a real-time image of the situation in emergency departments, helps to monitor gastrointestinal infection trends and informs public health measures. The timely surveillance of gastrointestinal infections in emergency departments facilitates outbreak detection and response. In summary, syndromic surveillance in emergency departments optimally complements statutory laboratory-based surveillance in Germany.

## 7. Data protection and ethical statement

The AKTIN emergency department registry received ethics approval from the Ethics Committee of the medical facility of the Otto von Guericke University in Magdeburg (ethics approval: 160/15 and 52/21). Primary data is stored (pseudonymized and decentralized) in local data warehouses at each hospital. AKTIN provides anonymized data to researchers after approval from their Data Use and Access Committee. Therefore, an additional ethics approval for this specific study is not required. Moreover, the use of routine emergency department data for surveillance was approved by the data protection officer at the Robert Koch Institute (BDS/ISB, 09-01-2019).

## 8. Data availability

Weekly aggregated data is available upon request by agreement with the AKTIN emergency department registry.

## 10. Acknowledgement

The authors would like to express their gratitude to the authors of the R packages *tidyverse*, *lubridate*, *ISOweek*, *Readxl*, *naniar*, *flextable*, *gtsummary*, *stringr*, *tsibble*, *slider*, *ggh4x*, *ggpubr* and *tseries*.

Moreover, the authors would like to thank the Postgraduate Training for Applied Epidemiology (PAE) and the European Programme for Intervention Epidemiology Training (EPIET) for enabling this study, particularly Sofie Gillesberg Raiser, Dr. Jan Walter and Dr. Raskit Lachmann.

## 12. Author’s contributions

JHJB, TSB and MS conceptualised the project. MS supervised the project. JHJB, MS and HW developed the syndrome definition. JHJB devised the study protocol, researched literature, performed data analysis, interpreted results, created visuals, and wrote the manuscript. MS, HW, TSB, provided feedback on the study protocol, methods, data analysis, and interpretation. AD assisted with the statistical analysis and KH helped with the literature research. SD, BE, RG, CG, KH, EH, AH, HH, CK, FR, JR, and TS gave clinical feedback on the syndrome definition. AKTIN Research Group provided the emergency department surveillance data used in this study. All authors critically reviewed the manuscript and approved the final version.

